# Harmonizing Palliative Care: National Survey to Evaluate the Knowledge and Attitude of Emergency Physicians towards Palliative Care

**DOI:** 10.1101/19003939

**Authors:** Ameena Mohammed Al-Ansari, Saleem Nawaf Suroor, Sobhi Mostafa AboSerea, Wafaa Mostafa Abd-El-Gawad

## Abstract

**Background and Aim:** Although the challenges of integrating palliative care practices across care settings are real and well recognized until now little is known about palliative care practice of emergency physicians (EPs) and their accessibility to palliative care services in Kuwait. So the aim of this study was to explore the attitude, and knowledge encountered by EPs in providing palliative care in all general hospitals in Kuwait.

**Method:** A cross-sectional survey was performed in the emergency rooms of all general hospitals in Kuwait using Palliative Care Attitude and Knowledge (PCAK) questionnaire.

**Results:** Of the total number of physicians working in emergency rooms (n=156), 104 (66.67%) had completed the survey. 76.9% (n=80) of the EPs had either uncertain attitude toward palliative care. Most of the EPs (n=73, 70.28%) didn’t discuss the need of the patients to palliative care either with the patients or their families. Only 16 (15.4%) of the EPs responded correctly to the most of the questions while nearly half of the EPs (n=51, 49%) had poor knowledge especially in the most effective management of refractory dysnea (n=18, 17.3%). Experience ≥ 11yrs and better knowledge scores were independent predictors of positive attitude after adjustment of age, sex, qualifications, specialty, position, and nationality [OR: 5.747 (CI: 1.031-25.00), 1.458(CI: 1.148-1.851); p-value: 0.021, 0.002 respectively]..

**Conclusions:** Despite recognizing palliative care as an important competence, the majority of the emergency physicians in Kuwait had uncertain attitude and poor knowledge towards palliative care. Lack of knowledge, direct accessibility to palliative care services and lack of support from palliative medicine specialists were the main reasons for uncertain and negative attitude. Efforts should be done to enhance physician training and provide palliative care resources in order to improve the quality of care given to patients visiting emergency departments.

**What this paper adds:**

- Studies proved that the emergency room may be a suitable place for early referral of patients who may benefit from palliative care especially old age to prevent upcoming undesired admissions and hospital deaths.
- The integration of palliative care concepts and consultation teams into emergency medicine may help to avoid unnecessary and burdensome treatments, tests, and procedures that are not aligned with patients’ goals of care.
- Although the challenges of integrating palliative care practices across care settings are real and well recognized until now little is known about palliative care practice of emergency physicians and their accessibility to palliative care services in Kuwait.
- Recently, a newly developed tool called Palliative Care Attitude and Knowledge (PCAK) questionnaire was created to assess the attitude and knowledge of non-palliative physicians toward palliative care. So the aim of this study was to explore the attitude, and knowledge encountered by emergency physicians in providing palliative care using PCAK ^8^ in emergency departments in all general
- Studies showed that early palliative care consultation was shown to improve quality of life for cancer patients and may even lengthen their survival.

**What this study adds:**

- Despite recognizing palliative care as an important competence, the majority of the emergency physicians in Kuwait had uncertain attitude and poor knowledge towards palliative care. Lack of knowledge, direct accessibility to palliative care services and lack of support from palliative medicine specialists were the main reasons for uncertain and negative attitude.
- Efforts should be done to enhance physician training and provide palliative care resources in order to improve the quality of care given to patients visiting emergency departments.

## Introduction

Emergency medicine is a medical specialty that is concerned with stabilization of patients with acute illness or injury for definitive care.^1^ in this specialty, emergency physicians are trained to provide acute treatments for emergency medical conditions aiming at preserving patients’ life regardless of their wellbeing.^1,2^

While patients and their families under palliative care can experience stressful and overwhelming moments during the disease trajectory such as sudden respiratory distress, severe pain, vomiting, confusion and many other symptom,^3^ the first point of access is often the emergency room. Unfortunately, many of them die in the hospitals despite their wish to die at home.^4^ Hospitals; where palliative medicine specialists are available; can provide the opportunity for the patients and their families to establish their preferences for care and to coordinate the aggressiveness of treatment, symptom management, and place of death.^3,4^

Moreover, early palliative care consultation was shown to improve quality of life for cancer patients and may even lengthen their survival.^5^ Most recently, studies proved that the emergency room may be a suitable place for early referral of patients who may benefit from palliative care ^6^ especially old age to prevent upcoming undesired admissions and hospital deaths.^7^

The integration of palliative care concepts and consultation teams into emergency medicine may help to avoid unnecessary and burdensome treatments, tests, and procedures that are not aligned with patients’ goals of care. ^2^ Although the challenges of integrating palliative care practices across care settings are real and well recognized until now little is known about palliative care practice of emergency physicians and their accessibility to palliative care services in Kuwait. Recently, a newly developed tool called Palliative Care Attitude and Knowledge (PCAK) questionnaire was created to assess the attitude and knowledge of non-palliative physicians toward palliative care.^8^ So the aim of this study was to explore the attitude, and knowledge encountered by emergency physicians in providing palliative care using PCAK ^8^ in emergency departments in all general hospitals in Kuwait.

## Method

### Study design and setting

A cross-sectional survey was performed in the emergency rooms of all general hospitals under the Ministry of Health of Kuwait. This includes six general hospitals; Al-Sabah, Al-Jahra, Al-Amiri, Al-Adan, Al-Farawanyia and Mubarak Al-Kabeer hospitals.

### Measurements and intervention

#### Palliative Care Attitude and Knowledge Questionnaire (PCAK):^8^

It is a newly developed questionnaire composed of three sections. Section one includes demographic data such as age, sex, level of education, work experience, workplace, medical subspecialty and palliative care experience. Section two is assessing the attitude and formed of 11 items. The tool had a 5 point Likert scale ranging from strongly disagree (1) to strongly agree (5). Negative or unfavorable attitude was considered if the participant scored =<25, uncertain attitude if scored >25 but <41, positive or favorable attitude if scored > =41. The third section inquiries about knowledge. It includes 2 parts; the first part was about the self-reported knowledge (3 questions) and the second part is 12 clinical questions. Regarding self-knowledge, 5 points likert scale was used ranging from excellent response (5) to none (1). Regarding basic knowledge scoring; Poor knowledge was calculated if participant scored less than 50% of the total score (12 points) (=<5 points), fair knowledge if >=50% to= <75% (6-9 points), good knowledge if scored >75% (>=10 points). ^8^

### Selection of Participants

The questionnaires were disturbed to all emergency physicians in all general hospital in Kuwait (Figure 1) and recollected back by hand to hand and revised within 48 hours. The flowchart of the sampling procedure was presented in Figure 1.

**Figure 1:**
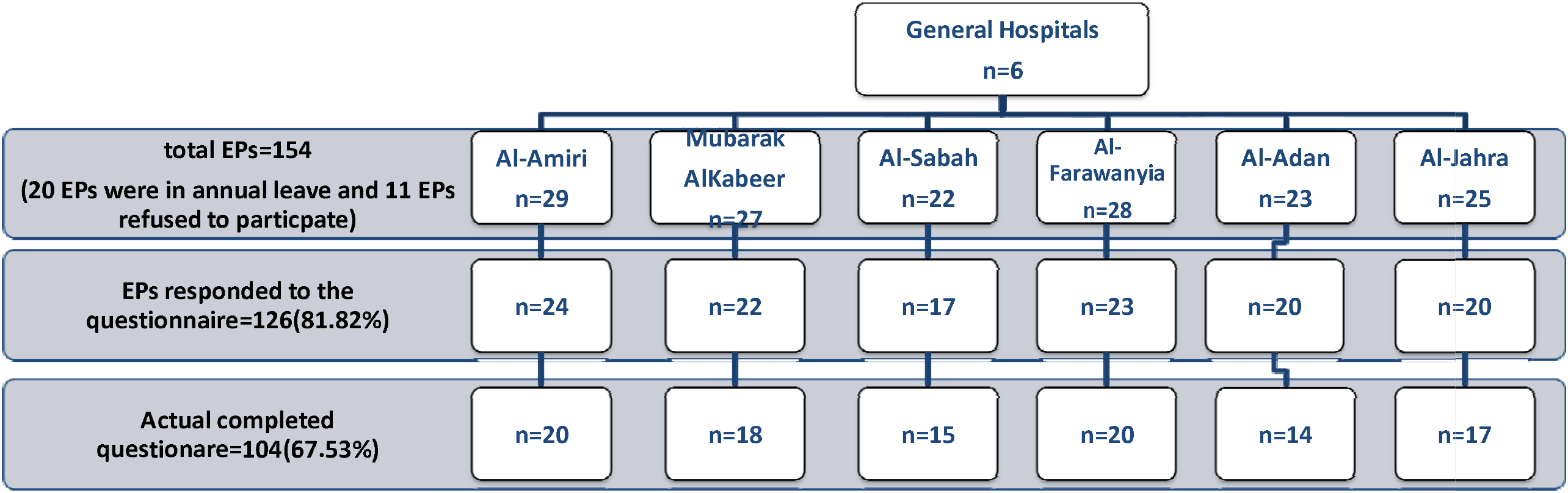
The Flowchart For the sampling procedure. EPs: emergency physicians

### Ethical statement

The approval of the Institutional Review Board (IRB) of the Ministry of Health was taken prior to the study (No.210/2016, March 2016). Informed consent was obtained from all participants. Participants’ anonymity and confidentiality were kept throughout all phases of the study. Approvals were also obtained from participating emergency departments of all general hospitals.

### Statistical analysis

All data manipulation and analysis were performed using the SPSS (Statistical Package for Social Science) SPSS version 20. P-values less than 0.05 were regarded as a sign of statistical significance. Independent sample t-test was used for comparison between quantitative data. Chi-square test or Fisher’s Exact when appropriate were used to compare between qualitative variables. Generalized linear method was used to find out the independent predictors of positive attitude toward palliative care with adjustment of any confounding factors. We used the consolidated criteria for reporting observational studies (RECORD) checklist as far as this was applicable to our study.

## Results

### Characteristics of study subjects

Of the total number of physicians working in emergency rooms in Kuwait (n=156 physicians), 19 physicians were on annual leave and 11 physicians refused to participate. The remaining 126 emergency physicians who responded to the questionnaire, 104 (82.54%) had completed the survey in all general hospitals. (Figure1). The number of participants was 15 (14.2%) from AlSabah hospital, 20 (19.2%) from Al-Amiri hospital, 18 (20%) from Mubarak Al-Kabeer hospital, 20 (17.33%) from Al-Farawanyia hospital, 14 (13.5%) from Al-Adan hospital and 17 (20.44%) from Al-Jahrah hospital. (Figure 1).

86.6% (n=90) were males and 13.5% (n=14) were females and mean age of the respondents was 38.83±9.5 years old. The median years of experience were 11 (IQR: 7-13) years. Due to the diversity of population demographics, only 20.19% (n=21) were Kuwaiti physicians. Most of respondents had postgraduate emergency medicine studies (n=68; 65.4%), followed by internal medicine studies (12; 11.5%). (Table 1).

**Table 1:** General description of the emergency physicians in Kuwait.

|  |  | Total<br>(N=104) | experience<br><11 yrs(n=50) | experience<br>≥11 yrs(n=54) | p-value* |
| --- | --- | --- | --- | --- | --- |
| Age |  | 38.83(±9.5) | 32.12(3.88) | 45.03(8.87) | <0.001 |
| Sex | Males | 90(86.6%) | 39(78%) | 51(94.4%) | 0.014 |
|  | females | 14(13.5%) | 11(22%) | 3(5.6%) |  |
| Nationality | Kuwaiti | 21(20.2%) | 18(38%) | 3(5.6%) | <0.001 |
|  | Non-Kuwaiti | 83(79.8%) | 32(64%) | 51(94.4%) |  |
| Qualification | MBBS | 30(28.8%) | 21(42%) | 9(16.7%) | 0.202** |
|  | Master | 59 (56.7%) | 24(48%) | 35(64.8%) |  |
|  | MD, MRCP/MRCS | 15(14.5%) | 5(10%) | 10(18.6%) |  |
| Specialty | ER | 68(65.4%) | 32(64%) | 36(66.7%) | 0.007** |
|  | Internal Medicine | 12(11.5%) | 2(4%) | 10(18.5%) |  |
|  | Surgery | 10(9.6%) | 4(8%) | 6(11.1%) |  |
|  | Family Medicine | 3(2.9%) | 3(6%) | 0 |  |
|  | Others | 11(10.6%) | 9(18%) | 2(3.7%) |  |
| Position | Assistant registrar | 6(5.8%) | 6(12%) | 0 | 0.020** |
|  | Registrar | 86(82.7%) | 40(80%) | 46(85.2%) |  |
|  | Senior registrar | 8(7.7%) | 4(8%) | 4(7.4%) |  |
|  | Specialist/ Consultant | 4(3.8%) | 0 | 4(7.4%) |  |
| Discussion about<br>palliative care | No patients | 73(70.2%) | 34(68%) | 39(72.2%) | 0.726** |
|  | 1 to 5 patients | 23(22.1%) | 11(22%) | 12(22.2%) |  |
|  | 6 to 10 patients | 6(5.8%) | 4(8%) | 2(3.7%) |  |
|  | 11 to 15 patients | 1(1%) | 1(2%) | 0 |  |
|  | > 15 patients, families | 1(1%) | 0 | 1(1.9%) |  |
| Self-assessment of his knowledge in: |  |  |  |  |  |
| 1-Pain | Excellent | 3(2.9%) | 1(2%) | 2(3.7%) | 0.311** |
|  | very good | 15(14.4%) | 8(16%) | 7(13%) |  |
|  | good | 42(40.4%) | 21(42%) | 21(38.9%) |  |
|  | weak | 29(27.9%) | 10(20%) | 19(35.2%) |  |
|  | none | 15(14.4%) | 10(20%) | 5(9.3%) |  |
| 2-Other Symptoms | Excellent | 4(3.8%) | 2(4%) | 2(3.7%) | 0.683** |
|  | very good | 21(20.2%) | 10(20%) | 11(20.4%) |  |
|  | good | 58(55.8%) | 28(56%) | 30(55.6%) |  |
|  | weak | 14(13.5%) | 5(10%) | 9(16.7%) |  |
|  | none | 7(6.7%) | 5(10%) | 2(3.7%) |  |
| 3-Councling | Excellent | 6(5.8%) | 2(4%) | 4(7.4%) | 0.402** |
|  | very good | 29(27.9%) | 15(30%) | 14(25.6%) |  |
|  | good | 41(39.4%) | 20(40%) | 21(38.9%) |  |
|  | weak | 18(17.3%) | 6(12%) | 12(22.2%) |  |
|  | none | 10(9.6%) | 7(14%) | 3(5.6%) |  |

\*p- value < 0.05 is significant.
\*\* Fisher Exact

**Table 2:**
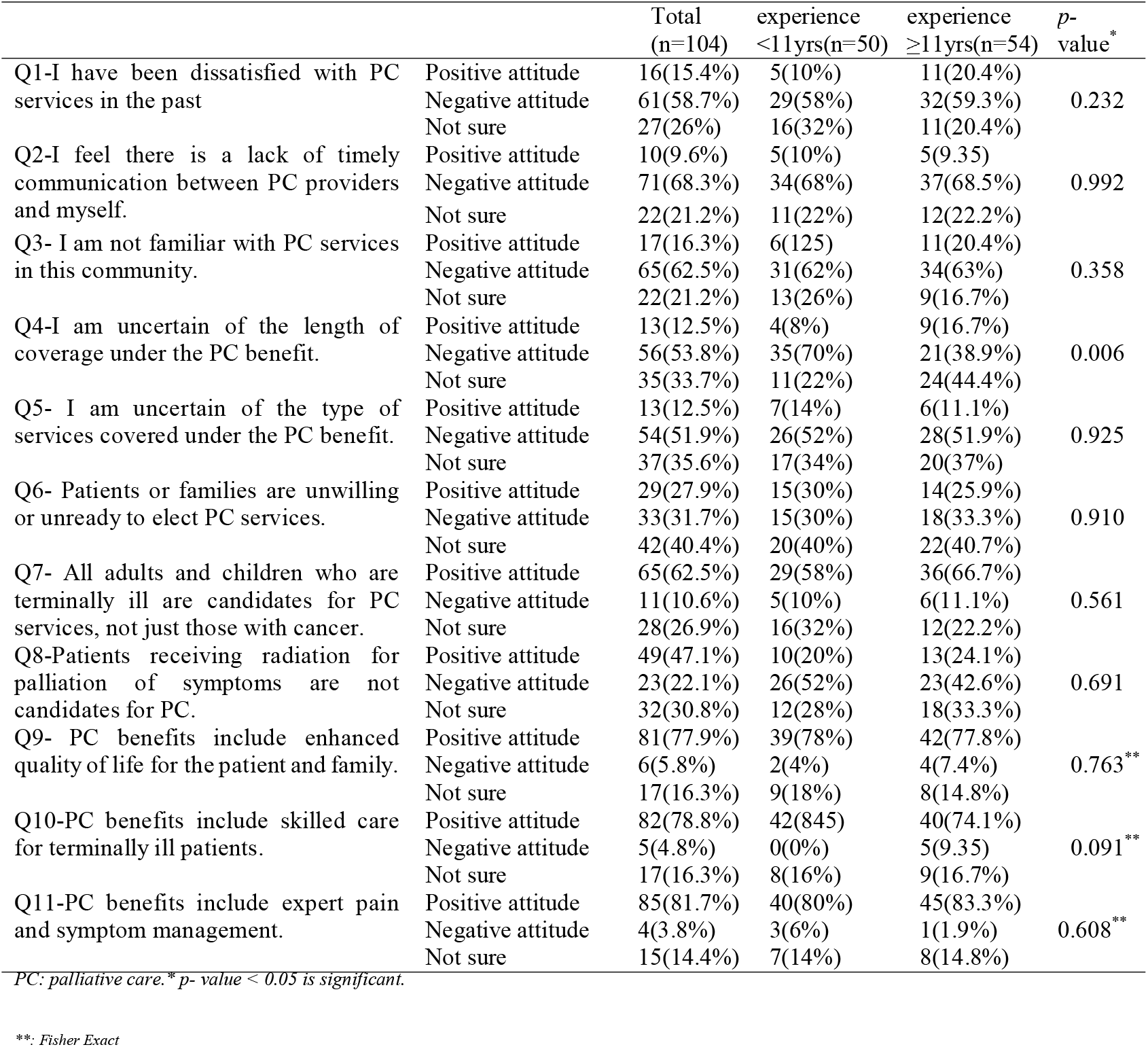
Distributions of the emergency physicians’ attitude towards palliative care in Kuwait.

### Main results

Only 18.3% (n=19) of the emergency physicians had a favorable attitude towards palliative care. Most of them had uncertain attitude (76.9%, n=80) and 4.8% (n=5) had an unfavorable attitude. Most of them agreed about the need of all adults and children who are terminally ill for palliative care services, not just those with cancer (Q7: n =65; 62.5%) and approved also that palliative care benefits include enhanced quality of life for the patient and family (Q9:n=81; 77.9%), skilled care for terminally ill patients (Q10: n=82; 78.8%), and expert pain and symptom management (Q11: n=85; 81.7%). Unfortunately, they emphasized onthe lack of timely communication between palliative care providers and themselves (Q2: n=71; 68.3%). Many of them dissatisfied with palliative care services in the past (Q1: n=61; 58.7%), reported unfamiliarity with the currently available palliative care services (Q3: n=65; 62.5%), and its types (Q5:n=54; 51.9%). Moreover, the uncertainty about its length of coverage was 53.8% (Q4: n=56). Generally, the emergency physicians with experience ≥11 years had statistically significant better attitude scores than the emergency physicians with experience <11 (p value=0.019, Figure 2) but in most of the individualized items of the attitude, no statistically significant between them.

**Figure (2):**
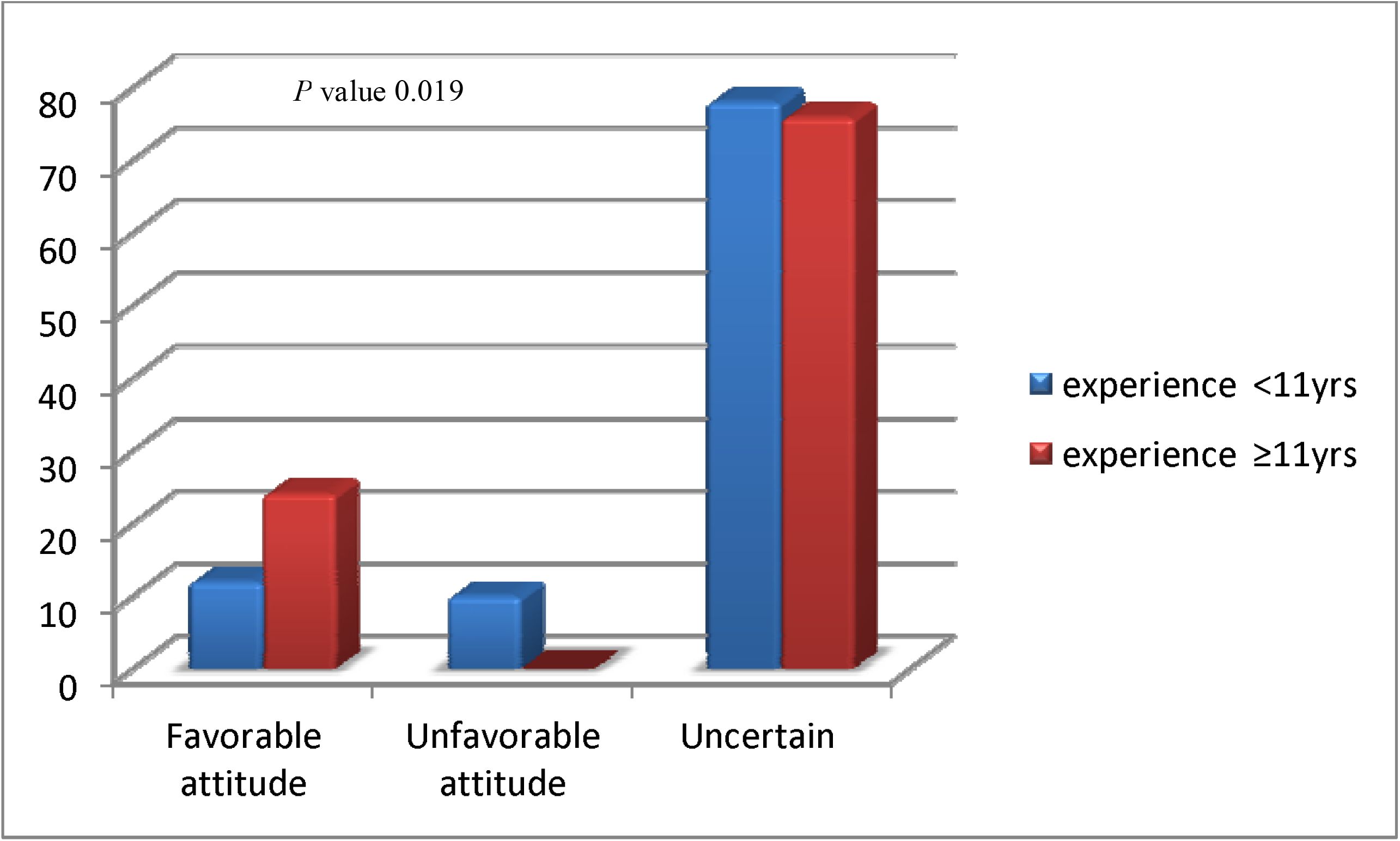
Attitude of Emergency physicians toward Palliative Care.

Most of the emergency physicians (n=73, 70.28%) didn’t discuss the need of the patients to palliative care either with the patients or their families. 44 (42.3%) of them reported no or little knowledge in pain assessment and management, while 58 (55.8%) and 41 (39.4%) reported good knowledge in other symptoms assessment and management, and the counseling respectively. Neither years of experience nor nationality had any statically significant effect on self-assessment of their knowledge and experience of the emergency physicians towards palliative care. (Table 1).

The overall percentage of the emergency physicians that responded correctly to the most of the questions about the basic knowledge of palliative care was only 16 (15.4%) while 37 (35.6%) had a fair knowledge and unfortunately many of them (n=51; 49%) responded to > 50% of the questions incorrectly. The median number of total corrected answers was 6 (IQR: 4-8) out of 12. Surprisingly, none of the emergency physicians answered all the questions correctly. An overview of the questions and their answers was presented in Table 3. Q11 for the symptoms of cord compression was the one with the highest percentage of correct answers (Q11: n=94; 90.4%) while Q8 about the most effective treatment for the refractory dyspnea question had the lowest rate of correct answers (Q8:n=18;17.3%). They had mostly poor to fair knowledge regarding opioids handling; such as types of opioids (Q3:n=57;54.8%), World Health Organization ladder for pain management (Q7: n=39;37.5%), opioids toxicity (Q8:n=31; 29.8%), use of oral opioids (Q15:n=19; 18.3%); and delirium (Q4:n=62; 59.65%) and management of catastrophic bleeding in palliative setting (Q10:n=20; 19.2%).

**Table 3:**
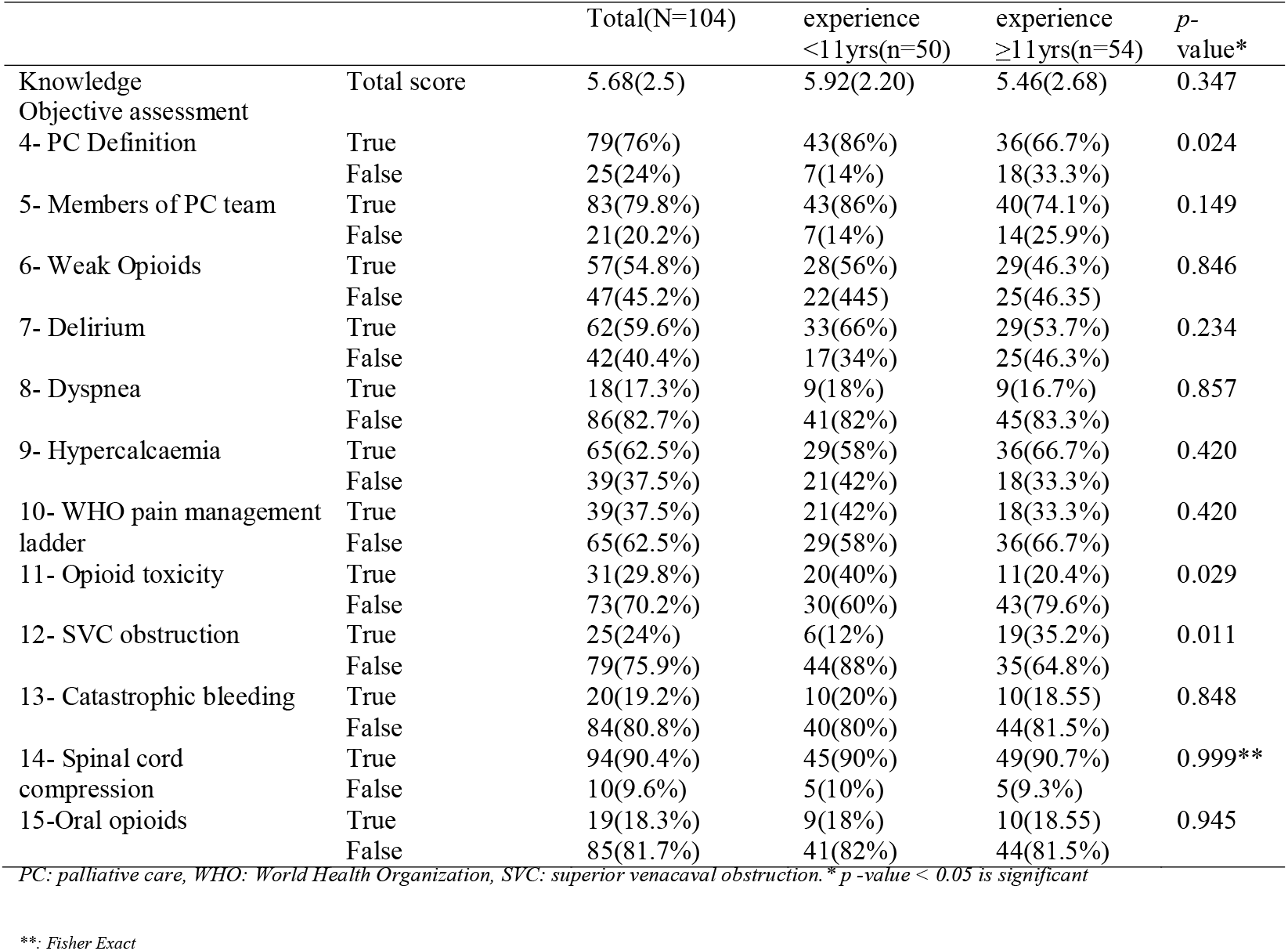
Distributions of the emergency physicians’ knowledge towards palliative care in Kuwait.

Most of the emergency physicians had good knowledge scores regarding the differences between traditional and palliative care (Q1: n=79; 76%) and the multidisciplinary team role in palliative care (Q2:n=83; 79.8%). No statistically significant differences between years of experience of the emergency physicians and their knowledge scores (Figure 3) in most of the questions except palliative care definitions and signs of opioid toxicity in which the emergency physicians with less experience answered significantly better (86% vs 66.7%, 40% vs 20.4%; p-value: 0.024, 0.029 respectively) while in diagnosis of superior venous obstruction, the emergency physicians with more years of experience answered better (12% vs 35.2%; p-value: 0.011). No statistically significant differences between the emergency physicians from different hospitals in the attitude or knowledge scores or demographic characteristics. (Table 4).

**Table 4:** Comparison between the emergency physicians’ characteristics working in different hospitals in Kuwait.

|  | Mubarak AlKabeer<br>(N=18) | AlAdan<br>(N=14) | AlSabah<br>(N=15) | Al-Jahra<br>(N=17) | Al-Farawanyia<br>(N=20) | Al-An<br>(N=20) |
| --- | --- | --- | --- | --- | --- | --- |
| Age | 40.11(±11.6) | 39.64(±10.4) | 43.11(±8.8) | 34.25(±5.2) | 39.4(±9.8) | 36.95(±10) |
| Sex |  |  |  |  |  |  |
| • Males | 14(77.8%) | 13(92.9%) | 14(93.3%) | 17(100.0%) | 16(80.0%) | 16(80.0%) |
| • Females | 4(22.2%) | 1(7.1%) | 1(6.7%) | 0 | 4(20.0%) | 4(20.0%) |
| Nationality |  |  |  |  |  |  |
| • Kuwaiti | 5(27.8%) | 2(14.3%) | 2(13.3%) | 1(5.9%) | 5(25.0%) | 6(30.0%) |
| • Others | 13(72.2%) | 12(85.7%) | 13(86.7%) | 16(94.1%) | 15(75.0%) | 14(70.0%) |
| Qualification |  |  |  |  |  |  |
| • MBBS | 4(22.2%) | 2(14.3%) | 1(6.7%) | 3(17.6%) | 7(35.0%) | 11(55.0%) |
| • Master | 12(66.7%) | 8(57.1%) | 8(53.3%) | 14(82.4%) | 12(60.0%) | 7(35.0%) |
| • MD | 2(11.1%) | 4(28.6%) | 6(40.0%) | 0 | 1(5.0%) | 2(10.0%) |
| Position |  |  |  |  |  |  |
| • Assistant registrar | 2(11.1%) | 0 | 0 | 1(5.0%) | 1(5.0%) | 2(10.0%) |
| • Registrar | 14(77.8%) | 10(71.4%) | 11(73.3%) | 16(94.1%) | 18(90.0%) | 17(85.0%) |
| • Senior registrar | 1(5.6%) | 4(28.6%) | 1(6.7%) | 0 | 1(5.0%) | 1(5.0%) |
| • Specialist/ Consultant | 1(5.6%) | 0 | 3(20%) | 0 | 0 | 0 |
| Specialty |  |  |  |  |  |  |
| • Emergency Medicine | 8(44.4%) | 12(85.7%) | 8(53.3%) | 10(58.8%) | 16(80.0%) | 14(70.0%) |
| • Internal Medicine | 5(27.8%) | 1(7.1%) | 2(13.3%) | 2(11.8%) | 1(5.0%) | 1(5.0%) |
| • Surgery | 0 | 0 | 3(20.0%) | 1(5.9%) | 3(15.0%) | 3(15.0%) |
| • Others | 5(27.8%) | 1(7.1%) | 2(13.3%) | 4(23.5%) | 0 | 2(10.0%) |
| Experience yrs) | 14.11(±9.6) | 14.79(±10.5) | 17.47(±8.5) | 9.38(±5.1) | 15.05(±10.3) | 10.9(±10.5) |
| Attitude |  |  |  |  |  |  |
| • Positive attitude | 7(38.9%) | 7(50.0%) | 5(33.3%) | 4(23.5%) | 5(25.0%) | 6(30.0%) |
| • Negative attitude | 2(11.1%) | 2(14.3%) | 4(26.7%) | 5(29.4%) | 3(15.0%) | 5(25.0%) |
| • Not sure | 9(50.0%) | 5(35.7%) | 6(40.0%) | 8(47.1%) | 12(60.0%) | 9(45.0%) |
| Knowledge |  |  |  |  |  |  |
| Total score | 4.78(2.25) | 6.0(2.51) | 6.07(2.81) | 6.12(2.48) | 5.65(2.43) | 5.6(2.43) |
| • Good knowledge | 1(5.6%) | 3(21.4%) | 4(26.7%) | 3(17.6%) | 3(15.0%) | 2(10.0%) |
| • Fair knowledge | 6(33.3%) | 4(28.6%) | 6(40.0%) | 7(41.2%) | 7(35.0%) | 7(35.0%) |
| • Poor knowledge | 11(61.1%) | 7(50.0%) | 5(33.3%) | 7(41.2%) | 10(50.0%) | 11(55.0%) |

\* P value < 0.05 is significant \*\*; Fisher Exact

**Figure (2):**
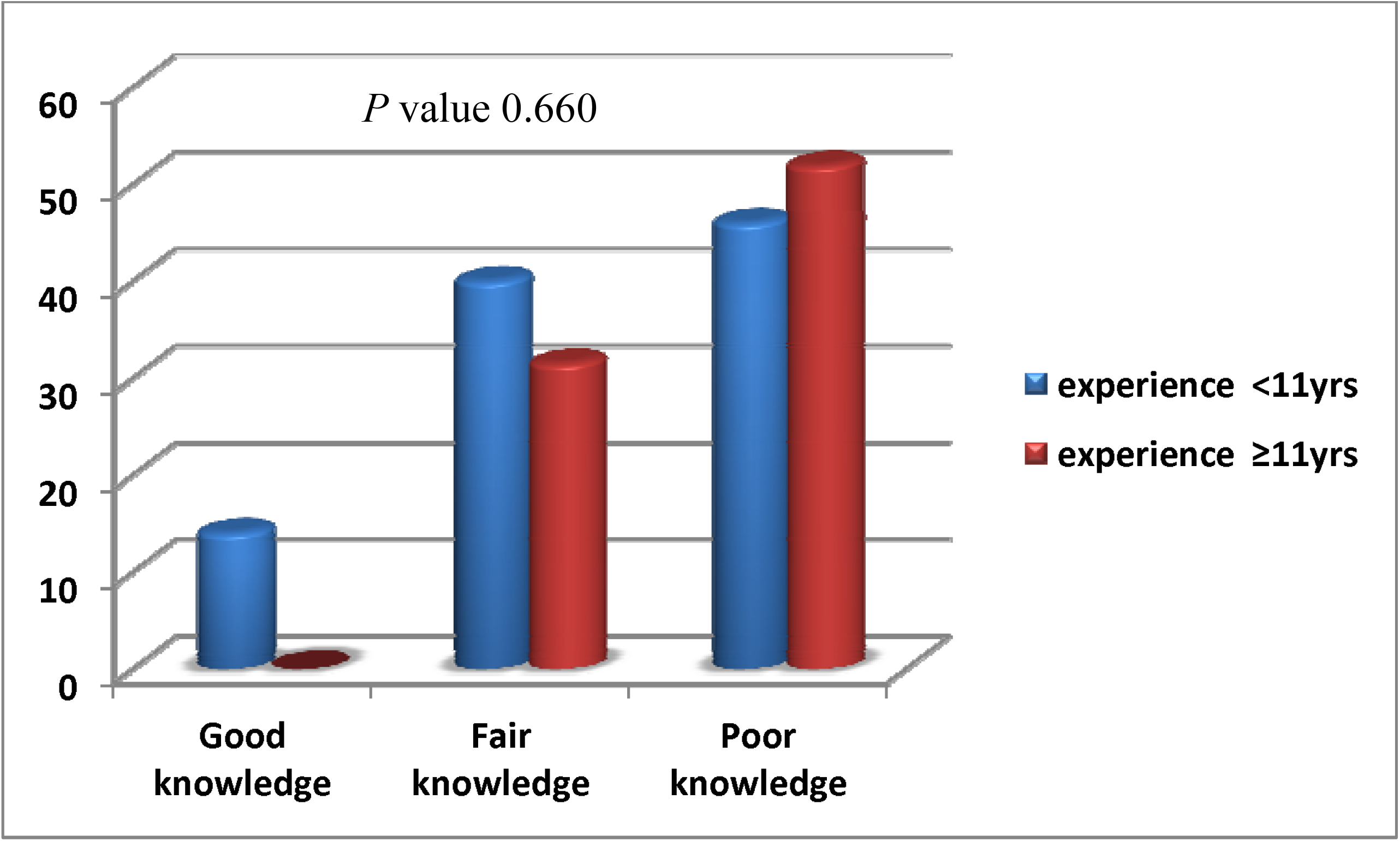
Knowledge of Emergency physicians toward Palliative Care.

By using generalized linear method to find out the independent predictors of positive attitude toward palliative care, we found that years of experience ≥ 11 yrs and better knowledge scores were independent predictors of positive attitude after adjustment of age, sex, qualifications, specialty, position, and nationality [OR: 5.747 (CI: 1.031-25.00), 1.458(CI: 1.148-1.851); p-value: 0.021, 0.002 respectively]. (Table 5)

**Table (5):** Generalized linear model of predictors of positive attitude:

|  | B | SE | OR | 95% CI |  | p- value |
| --- | --- | --- | --- | --- | --- | --- |
|  |  |  |  | Lower | Upper |  |
| Sex (male) | 0.641 | 0.805 | 1.899 | 0.392 | 9.194 | 0.426 |
| Age | 0.025 | 0.038 | 1.026 | 0.953 | 1.104 | 0.501 |
| Nationality (Kuwaiti) | 0.393 | 0.809 | 1.482 | 0.304 | 7.233 | 0.627 |
| Qualification (master, MBBS) | 0.621 | 1.01 | 0.538 | 0.075 | 3.877 | 0.538 |
| Position (assistant registrar or registrar) | 0.419 | 1.19 | 0.658 | 0.064 | 6.752 | 0.724 |
| Subspecialty | 0.399 | 0.567 | 0.538 | 0.075 | 3.877 | 0.538 |
| Years of experience | 1.75 | 0.756 | 5.747 | 1.031 | 25.00 | 0.021 |
| Basic knowledge score | 0.377 | 0.122 | 1.458 | 1.148 | 1.851 | 0.002 |

\* *p* -value < 0.05 is significant

## Discussion

To the best of our knowledge, this is the first survey of its kind conducted in Kuwait to evaluate palliative care attitude and knowledge targeting the emergency physicians anywhere in the entire region. We used PCAK which is a newly developed validated and reliable tool ^8^ to assess the attitude and knowledge among physicians. There are more than 150 physicians working in the emergency rooms in 6 hospitals distributed all over Kuwait. Most of them had uncertain attitude and poor knowledge scores. Better knowledge scores and experience ≥11 yrs were positive predictors of favorable attitude. Although more than 10 different nationalities are existing in Kuwait like Egyptians, Syrians, Pakistanis, Indians, and others from different cultures, emergency rooms demographic, medical education, specialties, years of experience but no statically significant differences between them in their knowledge or attitude toward palliative care. This differed from many studies which reported that clinician attitudes or knowledge were affected by nationality, physicians’ position, staff type, years of experience, emergency rooms demographic and hospital type.^9^

In our study, 76.9% of the emergency physicians had either uncertain attitude toward palliative care while only 18.3% had a favorable or positive attitude. This was mainly due to either dissatisfaction about PC services, types, accessibility, length of coverage and lastly lack of timely communication between them and palliative care providers as reported in our study. These results is similar to the original results of PCAK questionnaire.^8^ Many factors affecting the attitude of the emergency physicians depending on their diverse experiences, which ranged from positive, encouraging to a negative and distressing expiernce.^9,10^ This includes sometimes a sense of incompetence to alter the course of the disease ^3,11^ or feeling anxious and uncomfortable about discussing death and dying with the terminally ill patient.^12,13^ This negative attitude can hinder the quality of care provision especially at the end of life.

Most of the emergency physicians (n=73, 70.28%) didn’t discuss the need to palliative care either with the patients or their families. The willingness to engage in end-of-life discussions becomes key to improve palliative care in the emergency department.^14,15^ However, palliative care referrals remain largely underutilized in all areas of healthcare and are characteristically initiated late in the disease trajectory.^16,17^ Furthermore, most of the emergency physicians feel that it is not their obligation to discuss the need for palliative care and they feel that the family physicians or the oncologists are the one who is responsible about this discussion.^3,18^

Nearly half of the emergency physicians (49%) had poor knowledge. Although many of them subjectively reported good knowledge in counseling and other symptoms (rather than pain) assessment and management and they strongly agreed that palliative care is an important competence for emergency physicians ^3,18^ but it wasn’t necessarily reflected in their effectiveness of care especially in view of their poor knowledge at most. These results is similar to the original results of PCAK questionnaire.^8^

They had mostly poor knowledge in opioids handling, delirium and management of catastrophic bleeding in the palliative setting. This is similar to many studies that reported a lack of knowledge in palliative care, especially pain management proficiency.^18-20^

The setting of emergency rooms; as crowded spaces, noisy environment, compromised privacy, frequent interruptions, time constraints, illness complexity, and medico-legal threats; ^21,22^ make the initiation of palliative care discussions less than ideal in these settings. ^3,22^ Unfortunately, being not confident with the diagnosis and concerns of diminishing patients’ or families’ hope, or believing that patients are not prepared to hear forthcoming information make physicians often hesitate to discuss patients prognosis either with the patients or their families.^22^ Subsequently, aggressive interventions often initiated may ultimately be misaligned with overall goals of care although retrospectively viewed as futile, harmful, and painful.^23^ In reality, the culture of emergency setting that provides stabilization of acute medical emergencies was sometimes at odds with the culture of palliative care, which balances quality of life with the burdens of invasive treatments.^2,18^

In our study, good knowledge was positively associated with the positive attitude of the emergency physicians toward palliative care. This was agreed with Thulesius et al. who revealed that good knowledge of staff significantly improved attitudes toward the end of life care and the authors suggested initiatives to educate physicians working in ED’s will help them to change their negative attitudes and contribute better quality of patient care. ^24^

Many studies reported a significant correlation between the level of knowledge and attitudes towards palliative care. This is highlighting that as participants’ level of knowledge increased, attitudes become more positive in hospitals such as in Lebanon,^25^ Ethiopia,^26^ Saudi Arabia,^27^ and India.^28^ As a part of human nature, the degree and complexity of their knowledge affect their attitudes and in turn their behavior.^29^

The effective measurement of knowledge and attitude of the physicians is an important component of the evaluation of education and practice especially if used prior to any educational activity or program. This would result in more willingness to contribute in discussion and generate greater receptivity to the educational materials.^30,31^ Again it can be applied post activity to document the change in their knowledge and attitude and at the same time to evaluate the efficiency of the training and educational program.

### Strength and limitation of the study

It is the first study to assess the knowledge and attitude of the emergency physicians toward palliative care in the entire region. All the emergency physicians in Kuwait were involved in the research with no selection bias. The scarcity of similar studies carried out in Kuwait, Eastern Mediterranean region and in other parts of the world made the comparison and discussion difficult. Palliative care is recently introduced in Kuwait and it is still in the phase of development with the shortage of the number of the available palliative medicine specialists, this may explain the uncertain or the negative attitude towards palliative care.

Despite recognizing palliative care as an important competence, the majority of the emergency physicians in Kuwait had uncertain attitude and poor knowledge towards palliative care. Lack of knowledge, direct accessibility to palliative care services and lack of support from palliative medicine specialists were the main reasons for uncertain and negative attitude.

## Conclusion

Despite recognizing palliative care as an important competence, the majority of the emergency physicians in Kuwait had uncertain attitude and poor knowledge towards palliative care. Lack of knowledge, direct accessibility to palliative care services and lack of support from palliative medicine specialists were the main reasons for uncertain and negative attitude. Efforts should be done to enhance physician training and provide palliative care resources in order to improve the quality of care given to patients visiting emergency departments. Attention should be given to palliative care by the national health policy. There is an urgent need to be incorporated into the national curriculum of medical students and the emergency physicians’ education. Further efforts are needed to push the palliative care across different healthcare sectors.

### Ethics approval and consent to participate

The research project has been approved by the Institutional Review Board (IRB) of the Ministry of Health, Kuwait (No.210/2016, March 2016) within which the work was undertaken and that it conforms to the provisions of the Declaration of Helsinki. All subjects gave informed consent and their anonymity was preserved.

### Consent for publication

not applicable.

## Data Availability

data available with the corresponding author upon reasonable request

## Availability of data and materials

The datasets generated during and/or analyzed during the current study are available from the corresponding author on reasonable request.

## Authors ’ contributions

**Ameena Al-Ansari, Saleem Nawaf Suroor, Sobhi Mostafa AboSerea, Wafaa Mostafa AbdElGawad:** study concept, study design, acquisition of subjects and data, interpretation of data, preparation of the manuscript. All authors had approved the final article.

## Acknowledgements

The authors would like to thank all the participants for their valuable contributions to this study.

## References

1. Schneider SM1, Hamilton GC, Moyer P, Stapczynski JS. Definition of emergency medicine. Acad Emerg Med 1998;5(4):348–51.

2. Grudzen CR1, Richardson LD, Hopper SS, Ortiz JM, Whang C, Morrison RS. Does palliative care have a future in the emergency department? Discussions with attending emergency physicians. J Pain Symptom Manag. 2012;43(1):1–9.

3. Rivera MR, Torres FS. Physician Attitudes on the Provision of Palliative Care in Puerto Rican Emergency Departments. J Palliat Care Med 2015; 5: 201.

4. Pritchard RS, Fisher ES, Teno JM, Sharp SM, Reding DJ, et al. Influence of patient preferences and local health system characteristics on the place of death. SUPPORT Investigators. Study to understand prognoses and preferences for risks and outcomes of treatment. Journal of the American Geriatric Society 1998; 46: 1242–1250.

5. Temel JS, Greer JA, Muzikansky A, et al. Early palliative care for patients with metastatic non-small-cell lung cancer. N Engl J Med 2010; 363:733–742. [PubMed: 20818875]

6. Lipinski M, Eagles D, Fischer LM, Mielniczuk L, Stiell IG. Heart failure and palliative care in the emergency department. Emerg Med J 2018; 35(12):726-729. doi: 10.1136/emermed-2017-207186.

7. Beynon T, Gomes B, Murtagh FEM, Glucksman Ed, Parfitt A, Burman R, et al. How common are palliative care needs among older people who die in the emergency department? Emerg Med J 2011;28(6):491–5. doi: 10.1136/emj.2009.090019.

8. Al-Ansari AM, Suroor SN, AboSerea SM, Abd-El-Gawad WM. Development of palliative care attitude and knowledge (PCAK) questionnaire for physicians in Kuwait. BMC Palliat Care. 2019;18(1):49. doi:10.1186/s12904-019-0430-9. PubMed PMID: 31170968; PubMed Central PMCID: PMC6555752.

9. Cheung KY, Chan KC. Experiences of healthcare professionals in providing palliative end-of-life care to patients in emergency departments: a systematic review protocol. JBI Database System Rev Implement Rep 2016;14(10):9–14.

10. Jackson V, Mack J, Matsuyama R, Lakoma M, Sullivan A, Arnold R, et al. A qualitative study of oncologists’ approaches to end-of-life care. J Palliat Med 2008; 11(6):893–906.

11. Peters L, Cant R, Payne S, O’Connor M, McDermott F, Hood K, et al. How death anxiety impacts nurses’ caring for patients at the end of life: a review of literature. Open Nurs J 2013;7:14–21.

12. Tait V, Higgs M, Magann L, Dixon J, Davis J, Fernandez R. Attitudes of nonpalliative care nurses towards palliative care. Intl J Palliat Care 2015;2015:1–6.

13. Hui D, Bansal S, Park M, Reddy A, Cortes J, Fossella F, et al. Differences in attitudes and beliefs toward endof-life care between hematologic and solid tumor oncology specialists. Ann Oncol 2015;26(7): 1440–6.

14. Shearer FM, Rogers IR, Monterosso L, Ross-Adjie G, Rogers JR. Understanding emergency department staff needs and perceptions in the provision of palliative care. Emerg Med Australas 2014;26(3):249–55.

15. Heaston S, Beckstrand RL, Bond AE, Palmer SP. Emergency nurses’ perceptions of obstacles and supportive behaviors in end-of-life care. J Emerg Nurs 2006;32(6):477–85.

16. Kistler EA, Sean Morrison R, Richardson LD, Ortiz JM, Grudzen CR. Emergency department-triggered palliative care in advanced cancer: proof of concept. Acad Emerg Med 2015;22(2):237–9.

17. Zimmermann C, Swami N, Krzyzanowska M, Hannon B, Leighl N, Oza A, et al. Early palliative care for patients with advanced cancer: a cluster-randomised controlled trial. Lancet 2014;383(9930):1721–30.

18. Stone SC, Mohanty S, Grudzen CR, Shoenberger J, Asch S, Kubricek K, Lorenz KA Emergency medicine physicians’ perspectives of providing palliative care in an emergency department. J Palliat Med. 2011; 14(12):1333–8. doi: 10.1089/jpm.2011.0106.

19. Goldonowicz JM, Runyon MS, Bullard MJ. Palliative care in the emergency department: an educational investigation and intervention. BMC Palliative Care 2018; 17:43. https://doi.org/10.1186/s12904-018-0293-5

20. Chan GK. End-of-life models and emergency department care. Acad Emerg Med 2004;11(1):79–86.

21. Jesus J, Geiderman J, Venkat A, et al. Physician orders for life-sustaining treatment and emergency medicine: ethical considerations, legal issues, and emerging trends. Ann Emerg Med. 2014;64(2):140–4.

22. Limehouse WE, Feeser VR, Bookman KJ, Derse A. A model for emergency department end-of-life communications after acute devastating events—part II: moving from resuscitative to end-of-life or palliative treatment. Acad Emerg Med 2012;19(11):1300–8.

23. Forero R, McDonnell G, Gallego B, et al. A Literature Review on Care at the End-of-Life in the Emergency Department. Emergency Medicine International. 2012;2012:486516.

24. Thulesius H, Petersson C, Petersson K, Håkansson A. Learnercentred education in end-of-life care improved well being in home care staff: a prospective controlled study. Palliat Med 2002;16: 347–354.

25. Huijer AH, Dimassi H. Palliative care in Lebanon: knowledge, attitudes and practices of physicians and nurses. J Med Liban 2007;55(3):121–8.

26. Kassa H, Murugan R, Zewdu F, Hailu M, Woldeyohannes D. Assessment of knowledge, attitude and practice and associated factors towards palliative care among nurses working in selected hospitals, Addis Ababa, Ethiopia. BMC Palliat Care 2014;4:13(1):6.

27. Abudari G, Zahreddine H, Hazeim H, Assi MA, Emara S. Knowledge of and attitudes towards palliative care among multinational nurses in Saudi Arabia. Int J Palliat Nurs 2014;20(9):435–41.

28. Das A, Haseena T. Knowledge and Attitude of Staff Nurses Regarding Palliative Care. International Journal of Science and Research 2015; 4 (11):1790–5.

29. Fabrigar LR, Petty RE, Smith SM, Crites SL Jr. Understanding knowledge effects on attitude-behavior consistency: the role of relevance, complexity, and amount of knowledge. J Pers Soc Psychol 2006;90(4):556–77.

30. Kawaguchi S, Mirza R, Nissim R, & Ridley J. Internal Medicine Residents’ Beliefs, Attitudes, and Experiences Relating to Palliative Care: A Qualitative Study. American Journal of Hospice and Palliative Medicine 2017; 34(4), 366–372.

31. von Gunten C F, Twaddle M, Preodor M, Neely K J, Martinez J, & Lyons J. Evidence of improved knowledge and skills after an elective rotation in a hospice and palliative care program for internal medicine residents. Am J Hosp Palliat Care 2005;22(3):195–203.

